# Family-supervised disulfiram as a culturally grounded model for alcohol use disorder treatment in Sri Lanka: a pilot randomised controlled trial

**DOI:** 10.64898/2026.04.25.26350029

**Authors:** Mahesh Rajasuriya, Pubudu Chulasiri, Praneeth Ratnayake, David Plevin

**Affiliations:** Department of Psychiatry, University of Colombo, Sri Lanka; Riverland Mallee Coorong Local Health Network, Riverland General Hospital, Australia; Public Health Specialist, Ministry of Health, Sri Lanka; School of Biomedical Sciences, SLIIT Hemas Health Uni, Sri Lanka Institute of Information Technology, Malabe, Sri Lanka; School of Biomedical Sciences, Althia, SLIIT Hemas Health Uni, Malabe, Sri Lanka; Faculty of Humanities and Science, Sri Lanka Institute of Information Technology, Malabe, Sri Lanka; Ramsay Clinic Adelaide and Adelaide University

**Keywords:** Alcohol use disorder, Disulfiram, Pragmatic clinical trial, Randomised controlled trial, Addiction treatment, Sri Lanka

## Abstract

**Objectives:** To evaluate the effectiveness and cultural feasibility of family-supervised disulfiram as a first-line treatment for alcohol use disorder (AUD) in Sri Lanka, and to compare its clinical outcomes with standard therapy delivered at a tertiary psychiatric unit.

**Design:** Single-blind Randomized Controlled Trial known as ETAT-RCT (Efficacy of Two Alcohol Treatments) was conducted under routine clinical setup with three parallel groups: family-supervised disulfiram, locally developed psychosocial intervention, and routine treatment. Allocation was independently concealed; assessors were blinded. Analyses followed an intention-to-treat approach using repeated-measures ANOVA (group x time). This paper reports the disulfiram (test) versus routine treatment (control) comparison; the psychosocial intervention will be reported separately.

**Setting:** University Psychiatry Unit, National Hospital of Sri Lanka, Colombo (UPU, NHSLC).

**Participants:** Patients aged ≥14 years with AUD presenting to the unit were recruited consecutively without inducements. Planned allocation ratio was 1:1:1 with 31 participants per arm; key exclusions were lifetime psychotic disorder and current contraindication to disulfiram.

**Randomisation:** Participants were randomised into each treatment arm using an independent concealed paper-based allocation system.

**Intervention:** (1) family-supervised disulfiram, with psychoeducation/support only - DT arm, (2) a locally developed denormalization focused psychosocial programme - PT arm, and (3) standard therapy (motivational/cognitive/behavioural input; naltrexone permitted; no disulfiram/denormalisation) - ST arm.

**Outcome measures:** Primary outcome was Alcohol Use Disorders Identification Test (AUDIT) score at 12 months. Key secondary outcomes were past 30 day alcohol use via Timeline Follow-Back (TLFB); alcohol biomarkers [ALT (alanine aminotransferase), γ-GT (gamma-glutamyl transferase), MCV (mean corpuscular volume)]; locally developed measures of addiction-relevant cognitive, affective, behavioural factors [AARSU (Attitude Assessment Related to Substance Use), BARSU (Behaviour Assessment Related to Substance Use)]; and Quality of Life Enjoyment and Satisfaction Questionnaire Short Form (Q-LES-Q-SF). Outcomes were assessed at baseline, 6, and 12 months.

**Results:** Participants in DT (n=33) and ST (n=38) were comparable at baseline. Both groups showed clinically and statistically significant improvement in AUDIT scores over 12 months (DT: F=39.90, p<0.001; ST: F=49.90, p<0.001), with no group×time interaction (F<0.001, p=0.98). Biomarkers and AARSU, and BARSU and Q-LES-Q-SF to a lesser degree, mirrored the AUDIT pattern. TLFB did not change significantly over time in either arm (p>0.05). In moderator analyses, improvement in AUDIT was not moderated by baseline motivation (F=0.20, p=0.89) but was moderated by baseline AUD severity (F=7.70, p=0.007). No serious adverse events were attributed to disulfiram. Adherence to supervised dosing was generally high during periods of supervision but intermittent overall.

**Conclusions:** In this pilot RCT, family-supervised disulfiram achieved 12-month outcomes comparable to standard therapy in a tertiary Sri Lankan setting. Improvements were independent of baseline motivation and varied by baseline AUD severity. These findings may support family-supervised disulfiram as a culturally feasible first-line option in Sri Lanka; larger, adequately powered multicentre trials are warranted to confirm effectiveness and scalability.

**Trial registration:** **S**LCTR/2014/021

**Strengths and limitations of this study:**

- This pragmatic randomised controlled trial demonstrates an improved real world applicability and validity as it was conducted in an unmodified public-sector psychiatric setting.
- Strong generalisability of the study with similar health systems due to broad eligibility criteria of patients warranted the inclusion of regular and general patient cohort with alcohol use disorders, strengthening generalisability within similar health systems.
- Interventions were carried out without additional staff or patient monitoring reflecting routine clinical practice.
- Comprehensive assessment beyond abstinence alone with multidimensional outcomes such as alcohol related harm, biomarkers, cognitive behavioural change and quality of life.
- Minor potential in performance bias due to the nature of intervention where blinding study subjects and clinicians is not feasible.
- Sampling bias towards males and variability within the ST arm can affect the generalisability.

## Introduction

The Global Burden of Disease Study 2021 attributed 1.8 million annual deaths worldwide to alcohol use, predominantly among males (1). Approximately 400 million people aged 15+ years (7% of the global population) live with alcohol use disorder (AUD), just over half of them with severe AUD or dependence (2). The prevalence of severe AUD in Sri Lanka is slightly lower, at 2.5%, but the striking contrast is that pathological drinking in Sri Lanka seems to be almost exclusively limited to men (3). Among males, the prevalence of “hazardous drinking” was 5.2% and among females 0.02%; this may be partly explained by alcohol use by women being a social taboo in Sri Lanka (3).

While many pharmacological options are available to treat AUD, disulfiram was the first to be approved by the US Food and Drug Administration for AUD in 1949 (4). Disulfiram irreversibly inhibits aldehyde dehydrogenase, causing acetaldehyde accumulation and a deterrent aversive reaction called disulfiram-ethanol reaction (DER) upon alcohol consumption (5). Its efficacy depends on patient’s awareness of DER risks, suggesting a psychological component to this biological intervention (6).

Notably, supervised disulfiram therapy outperforms unsupervised use, which does not differ from placebo in efficacy (4,7). Family supervision of disulfiram treatment adherence is reported to be effective (8–11). Disulfiram remains relevant after 75 years of clinical use (4–7,12–15), especially in low-resource settings due to its affordability and primary care applicability (5). However, current therapeutic guidelines do not recommend disulfiram as a first line treatment for AUD (16–19), and it remains underutilised (17,20).

Supervised disulfiram, especially family-supervised, as a first line treatment for AUD may prove effective, especially in resource poor settings in the developing world where family involvement in culturally permitted. Sri Lanka is a middle-income country (3), with a social structure facilitating family involvement in medical care (21).

Assessment of family-supervised disulfiram as an AUD treatment in Sri Lanka calls for evaluation of effectiveness of such therapy in the real world rather than of its mere efficacy under highly controlled conditions. Pragmatic trials evaluate interventions under real-world conditions, prioritising external validity (22). It is also important to note that traditional AUD trials focus narrowly on alcohol consumption. However, broader and clinically relevant outcomes such as alcohol-related beliefs and behaviour and quality of life better reflect patient-centred recovery (23).

This paper presents the findings of a pragmatic RCT, titled Efficacy of Two Alcohol Treatments (ETAT; ETAT-RCT), comparing the effectiveness of family-supervised disulfiram therapy to standard treatment for AUD as practiced at the University Psychiatry Unit (UPU), National Hospital of Sri Lanka, Colombo (NHSLC). Therefore, this trial was designed to inform and improve routine clinical decision making for the first line AUD treatment in psychiatric services in Sri Lanka.

## Methods

### Trial design

The Efficacy of Two Alcohol Treatments (ETAT-RCT) was a pragmatic, randomised, single-blind trial conducted at the University Psychiatry Unit (UPU), National Hospital of Sri Lanka, Colombo (NHSLC). We compared supervised disulfiram therapy (DT) and psychosocial treatment (PT) against standard treatment (ST), with outcome assessors blinded to allocation. This paper reports DT versus ST; PT outcomes will be published separately. The trial followed CONSORT 2010 guidelines and was prospectively registered (SLCTR/2014/021). Ethics approval was granted by the University of Colombo (EC-13-65).

### Participants

Eligible participants were patients aged ≥14 years presenting to the UPU, NHSLC with alcohol use disorders (AUD). We employed passive recruitment (no active inducement) and confirmed diagnoses using either ICD-10 or DSM-IV criteria (whichever provided broader inclusion). Participants with current or past psychotic disorders (assessed via Mini International Neuropsychiatric Interview [MINI]), any disulfiram contraindications, and those who fail to meet MINI-confirmed AUD criteria were excluded. The MINI served dual purposes: excluding psychotic disorders while verifying AUD diagnoses.

### Sample size

The study compared two treatment groups (DT, ST) across three assessment points: Baseline (BAx), 6-month (6Ax) and 12-month (12Ax) follow-ups. Our primary analysis used a 2×3 repeated-measures ANOVA to test the interaction between group and time on alcohol consumption (measured via TimeLine Follow Back, TLFB). Power calculation assumed a medium effect size (f = 0.25, Cohen’s terminology), a conservative correlation (r = 0.1) between repeated measures, and appropriate power (α = 0.05, power = 0.80). The effect size was assumed considering the smallest difference that would justify a change in routine clinical practice. This yielded a required sample size of 31 per group (total N=93). For the current DT vs ST comparison (presented here), the relevant sample is 62 participants (31 per group).

### Recruitment Process

Eligibility assessment occurred in three sequential steps executed by a psychiatry senior registrar (PSR) prior to obtaining fully informed written consent: In the first stage of initial clinical screening, the PSR evaluated potential participants for AUD diagnosis (meeting either ICD-10 [harmful use/dependence] or DSM-IV [abuse/dependence] criteria), psychiatric comorbidities (particularly lifetime psychotic disorders), and stage of change (motivational readiness). Motivational readiness was clinically evaluated and recorded using the Prochaska and DiClemente’s stages of change model (24). Precontemplation and contemplation stages were categorised as low motivation while other stages were categorised as high motivation. In the second stage of structured diagnostic confirmation, the PSR administered the Sinhala-translated MINI (English Version 5.0.0) (25) to confirm AUD diagnosis and rule out lifetime psychotic disorders. In the third stage of final eligibility verification, the PSR applied the previously mentioned inclusion/exclusion checklist.

### Randomisation and Allocation Concealment

We implemented a rigorously concealed allocation system to prevent selection bias:

1. *Sequence Generation*

The randomisation sequence was computer-generated with 1:1:1 allocation to DT/PT/ST groups.

1. *Allocation Preparation*

An independent non-academic staff member prepared sealed allocation packs consisting of cards pre-printed with treatment assignment and a unique participant number sealed in a sequentially numbered, opaque envelope. There was a carbon paper lining on top of the card (to transfer participant details to the card without having to open the envelope) and aluminium foil underneath the card (to light-proof preventing any potential attempt to pre-empt the allocation).

During the recruitment period of participants, 124 patients presenting to the UPU were screened for eligibility based on the inclusion and exclusion criteria. Of these, 110 (88.9%) met inclusion criteria, and only 2 (<0.02%) declined participation due to undisclosed reasons, ending up with 108 selected participants being randomized.

### Blinding

Blinding of participants and treating healthcare staff was not feasible given the nature of the interventions which reflects routine clinical practice. The assessors of the outcome measures were blinded to the treatment group allocation as they only had to apply the assessment tools without obtaining any history from the participants.

### Interventions (AUD treatments) Organisation and delivery and follow up

1. *Supervised Disulfiram Therapy (DT)*

Disulfiram was administered with a novel protocol characterised by daily supervision of tablet ingestion by a family member (rather than the clinical team) and formal commitment to treatment adherence in the participant, which was achieved through participant and family member signing a triplicate declaration form (clinic record, patient copy, family copy) explicitly transferring primary responsibility for adherence to the patient. Dose of disulfiram was adjusted clinically within the range of 250–375 mg/day. Patients could voluntarily stop disulfiram and resume alcohol use and later restart treatment, while family members encouraged (but did not force) treatment re-initiation. The ETAT-RCT provided disulfiram (daily cost LKR 14 – 28, USD 0.07 – 0.15, at the time) free to the participant.

*2. Psychosocial Treatment (PT)*

A unique, locally developed framework distinct from conventional AUD psychosocial therapies was delivered by trained counselors at the UPU. No AUD medications were used. The distinctive difference of this approach to traditional psychosocial therapies was its strong focus on reversal of normalisation of alcohol and its use, i.e., denormalisation. Denormalisation (not prohibition) of alcohol use and dealcoholisation of social events and interpersonal interactions, deconstruction of expectation of positive effects from acute alcohol use and increasing the awareness about how alcohol use is transformed into a glamorous behaviour are the main strategies of this treatment.

*3. Standard Treatment (ST)*

The usual treatment delivered by the UPU consisted of many components, most of which all patients received, such as regular medical review, supportive psychotherapy, motivational interviewing, and CBT for alcohol use. However, only ∼30% of patients received naltrexone (50 mg/day) due to cost barriers. Cost of naltrexone (LKR 289 or USD 1.53/ day at the time) was borne by the participant hence prescribed only to those who could afford it. This reflects the usual practice at the UPU,

NHSLC. Frequency of contact with treating team matched in all groups. The ST arm reflected the routine practice at the UPU, including variability in access to naltrexone based on patient affordability

### Treatment fidelity

Consistent with our pragmatic trial design, the study intentionally avoided interventions to enhance adherence, such as adherence prompts and active monitoring. Participants received no reminders (e.g., calls, texts) to attend clinics or take medications. Clinicians did not implement pill counts, biochemical verification, or other adherence tracking beyond standard clinical documentation. No additional allocation of staff, patient monitoring, or follow up resources beyond routine outpatient care was introduced as part of the DT intervention.

### Outcome measurements

The efficacy of treatment was assessed through the improvement observed in current pathological alcohol use, i.e., the primary outcome. Outcome measures were carefully selected to reflect specific domains relevant to clinicians, policy makers, patients and their family which included changes in alcohol related harm, attitudes, behaviours and quality of life, rather than abstinence alone. The quantity (standard drinks/day in the previous month) and the quality (drinking patterns and consequences in the previous year) of alcohol consumption were measured through standardised interviewer-administered tools TLFB (26) and Alcohol Disorder Identification Test (AUDIT) – validated Sinhala version (27) respectively. This self-reported data was corroborated by the levels of indirect biomarkers, i.e., alanine aminotransferase (ALT), gamma-glutamyl transferase (γ-GT), and mean corpuscular volume (MCV). TLFB and biomarkers were administered at the recruitment (BAx-baseline assessment), at 6 months into therapy (6Ax-6-month assessment), and at 12 months into therapy (12Ax-12-month assessment). AUDIT was administered at BAx and 12Ax only as the recall period was 12 months. Treatment fidelity patterns were observed through case notes.

Secondary outcomes were the improvements observed in cognitive-affective-behavioural factors (AUD-related attitudes, beliefs, and behaviours) and functioning (quality of life, health status, and psychosocial functioning). Two validated, locally developed instruments (please see supplementary data), Attitude Assessment Related to Substance Use (AARSU) and Behaviour Assessment Related to Substance Use (BARSU), were administered by a trained interviewer at BAx, 6Ax, and 12Ax.

AARSU assessed maladaptive beliefs and motivational states (direct/indirect alcohol-related cognitions), while BARSU assessed alcohol-use behaviours and adaptive/maladaptive coping strategies. Each instrument consists of 15 agree/ disagree Likert-type statements and a higher scored indicates a positive AUD outcome. Quality of life, health status, and psychosocial functioning were evaluated using Quality of Life Enjoyment and Satisfaction Questionnaire – Short Form (Q-LES-Q-SF), a tool validated for substance users (28) administered by a trained interviewer at BAx, 6Ax, and 12Ax. Q-LES-Q-SF assessed physical health, mood, work/school functioning, social relationships, and general life satisfaction, and a higher score indicates greater satisfaction.

### Analysis Primary analysis

The primary outcome was analysed using a **two-way mixed ANOVA** with:

- **Between-subjects factor**: Treatment group (DT vs. PT vs. ST)
- **Within-subjects factor**: Time (BAx vs. 6Ax vs. 12Ax)
- **Dependent variable**: TLFB-measured alcohol consumption (standard drinks/month)

AUDIT scores, biomarkers, and the secondary outcomes (AARSU, BARSU, and Q-LES-Q-SF) were analysed identically to maintain consistency.

To evaluate whether the baseline motivation to change behaviour or the diagnosis influenced the final outcome, we analysed the combined sample (DT + ST) comparing high- versus low-motivation and less- versus more-severe AUD participants using the changes in AUDIT scores at the end of treatment.

### Intention-to-treat analysis

All participants randomised in the trial (N=108) were retained in the final analysis irrespective of their adherence to treatment protocols or follow-up attendance. To optimise data completeness, participants who did not attend scheduled clinic visits for their 6-month (6Ax) or 12-month (12Ax) assessments were contacted by telephone. Where in-person attendance was not feasible, interviewer-administered instruments (AUDIT, TLFB, AARSU, BARSU, and Q-LES-Q-SF) were completed via telephone.

This approach minimised missing data for interview-based measures, though biomarker collection (ALT, γ-GT, MCV) remained vulnerable to higher missingness rates due to the necessity of in-person testing.

### Statistical analysis

Missing continuous data was addressed using mean imputation in IBM SPSS Statistics for Windows, version 25 (IBM Corp., Armonk, N.Y., USA). While recognising that mean imputation lacks a formal framework for handling dependencies in repeated-measures designs and may consequently underestimate variance we selected this pragmatic approach for its interpretability and common usage in clinical trials. To evaluate the robustness of our findings, we employed an empirical strategy whereby mixed-design ANOVA analyses using the general linear model (GLM) procedure were conducted on the imputed dataset with sensitivity analyses comparing these results to complete-case scenarios.

## Results

### Participant flow

Overview of participant recruitment, randomization, allocation, follow-up, and analysis in each study arm is illustrated in the Figure 1.

**Figure 1.**
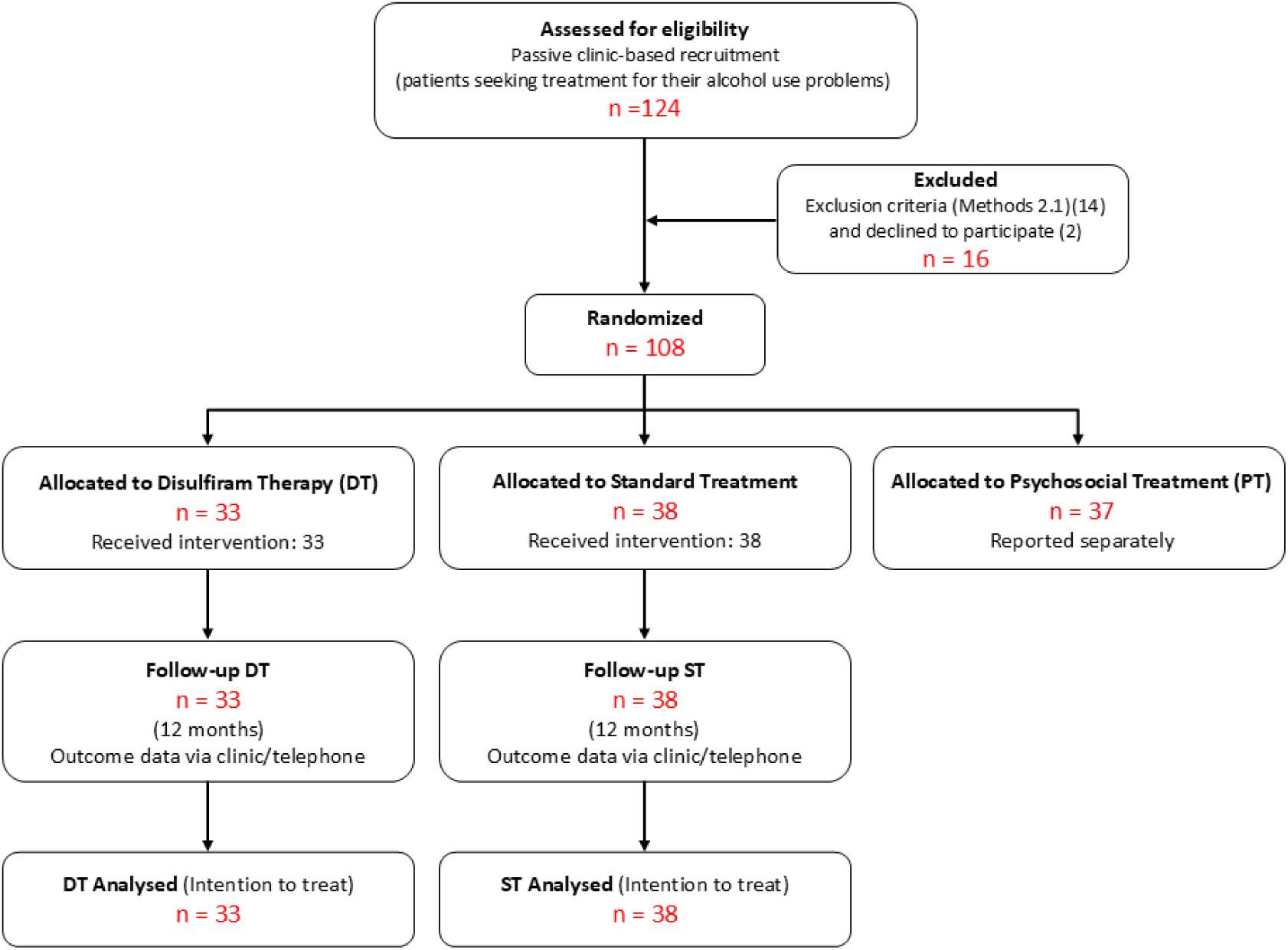
CONSORT diagram of the flow of study participants. Overview of participant recruitment, randomization, allocation, follow-up, and analysis in each study arm. Although one participant was deceased all participants were included in ITT analysis using statistical imputation

### Pragmatism

Evaluation of the ETAT-RCT methodology demonstrates a strong alignment with real-world clinical practice. As shown in Box 1, it satisfies seven of Ford and Norrie’s nine pragmatic trial dimensions (22).

Organisation and flexibility in delivery of treatment interventions were moderately controlled in the ETAT-RCT study. Hence, those two dimensions appear to have not been fully adhered to in the conduct of the study.

### Sample at the baseline

The study analysed 71 participants, comprising 33 in the disulfiram therapy (DT) group and 38 in the standard treatment (ST) group (see Table 1). Participants had a mean age of 46.9 years and were exclusively male, reflecting the gender distribution typical of alcohol use disorder presentations in this clinical setting. The sample was predominantly Sinhalese (74.6%, n=53) and Buddhist (70.4%, n=50), while most participants were married (63.4%, n=45) and had attained only a primary-level education (60.6%, n=43). Clinically, 93% (n=66) met diagnostic criteria for alcohol dependence, while only 36.6% (n=27) demonstrated high motivation for behavioural change at baseline, as assessed through standardized clinical evaluation.

**Table 1:** Characteristics of the sample (DT and ST groups)

| **Table 1: Characteristics of the sample (DT and ST groups)** |  |  |
| --- | --- | --- |
| **Sociodemographic** | **DT group**<br>**n (%)** | **ST group**<br>**n (%)** |
| Participants | 33 | 38 |
| Mean age | 45.9 | 47.8 |
| Male participants | 33 (100.0%) | 38 (100.0%) |
| Sinhala participants | 24 (72.7%) | 29 (76.3%) |
| Buddhist participants | 22 (66.7%) | 28 (73.7%) |
| Married participants | 21 (63.6%) | 24 (63.2%) |
| Participants that completed primary education only | 17 (51.5%) | 26 (68.4%) |
| Participants with alcohol dependence | 30 (90.9%) | 36 (94.7%) |
| Participants showing high motivation | 12 (36.4%) | 15 (39.5%) |
| DT group: Disulfiram therapy group |  |  |
| ST group: Standard therapy group |  |  |

To verify that the randomisation produced balanced treatment groups, we conducted chi-square tests comparing baseline characteristics between the DT and ST groups using BAx data. As presented in Table 2, the groups demonstrated comparable distributions across all measured demographic variables: age (χ²[df]= 0.341[1], p=0.59), ethnicity (χ²[df]=0.012[1], p=0.73), religion (χ²[df]=0.418[1], p=0.52), marital status (χ²[df]=3.72[2], p=0.15), and education level (χ²[df]=4.81[2], p=0.90). This confirms the absence of significant baseline differences between intervention arms prior to treatment initiation.

**Table 2:**
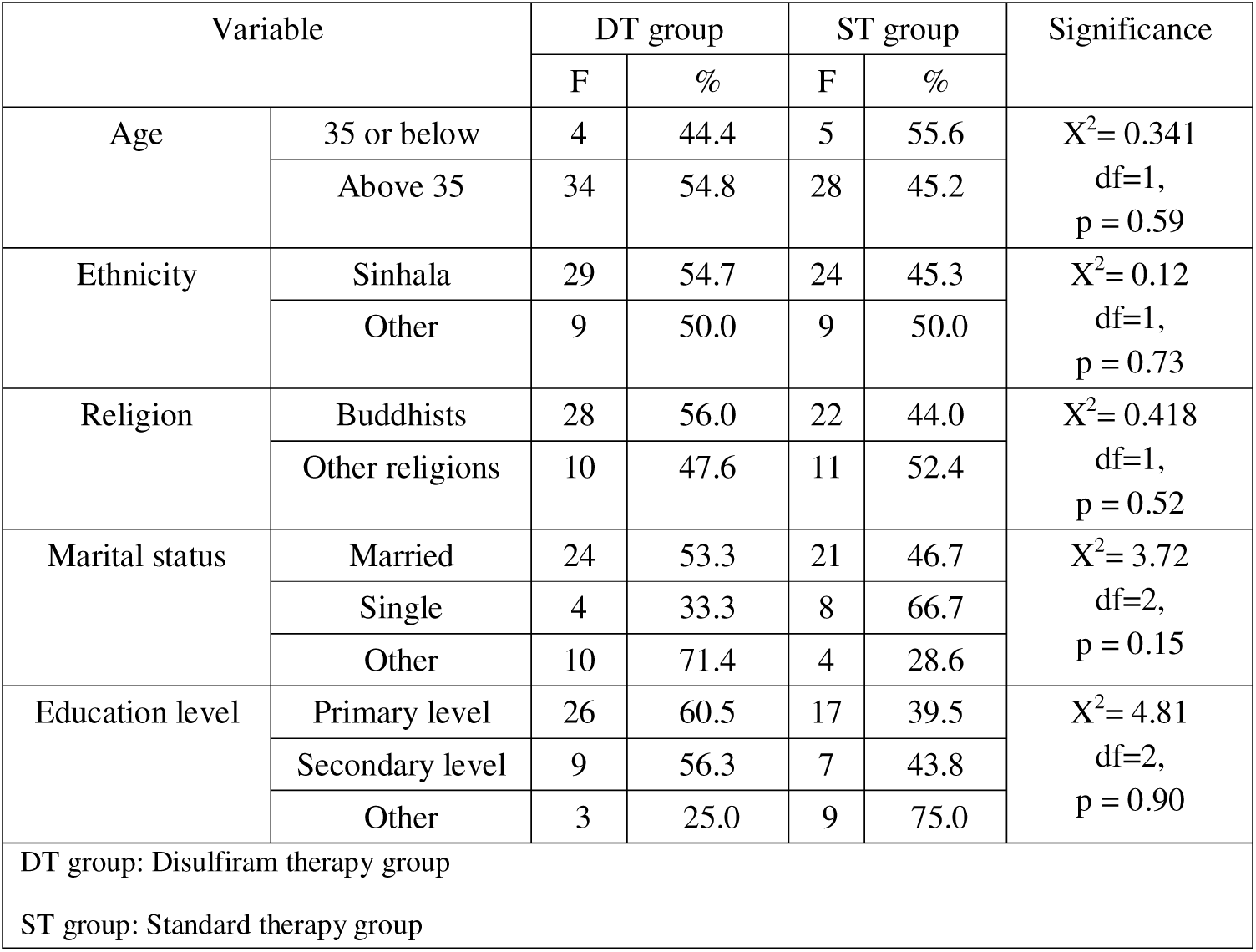
Comparison of the treatment (DT) and the control (ST) group at the baseline.

|  |  |  |  |  |  |  |
| --- | --- | --- | --- | --- | --- | --- |
| Variable |  | DT group |  | ST group |  | Significance |
|  |  | F | % | F | % |  |
| Age | 35 or below | 4 | 44.4 | 5 | 55.6 | X<sup>2</sup>= 0.341<br>df=1,<br>p = 0.59 |
|  | Above 35 | 34 | 54.8 | 28 | 45.2 |  |
| Ethnicity | Sinhala | 29 | 54.7 | 24 | 45.3 | X<sup>2</sup>= 0.12<br>df=1,<br>p = 0.73 |
|  | Other | 9 | 50.0 | 9 | 50.0 |  |
| Religion | Buddhists | 28 | 56.0 | 22 | 44.0 | X<sup>2</sup>= 0.418<br>df=1,<br>p = 0.52 |
|  | Other religions | 10 | 47.6 | 11 | 52.4 |  |
| Marital status | Married | 24 | 53.3 | 21 | 46.7 | X<sup>2</sup>= 3.72<br>df=2,<br>p = 0.15 |
|  | Single | 4 | 33.3 | 8 | 66.7 |  |
|  | Other | 10 | 71.4 | 4 | 28.6 |  |
| Education level | Primary level | 26 | 60.5 | 17 | 39.5 | X<sup>2</sup>= 4.81<br>df=2,<br>p = 0.90 |
|  | Secondary level | 9 | 56.3 | 7 | 43.8 |  |
|  | Other | 3 | 25.0 | 9 | 75.0 |  |
| DT group: Disulfiram therapy group |  |  |  |  |  |  |
| ST group: Standard therapy group |  |  |  |  |  |  |

### DT performance against ST

TLFB scores modestly reduced over time as shown in Table 3, but they failed to meet statistical significance in both DT (F=1.70, p=0.18) and ST (F=0.09, p=0.91) arms. This implies neither intervention produced statistically significant reductions in alcohol consumption during the study period. In contrast to TLFB scores, both DT (F=39.90, p=0.001) and ST (F=49.90, p=0.001) treatment groups demonstrated clinically and statistically significant improvements in AUDIT scores over the 12-month study period. Despite these substantial within-group reductions, between-group comparisons revealed no significant difference in efficacy (F<0.001, p=0.980), suggesting the there is no demonstrable superiority of DT approach compared to ST approach.

**Table 3:**
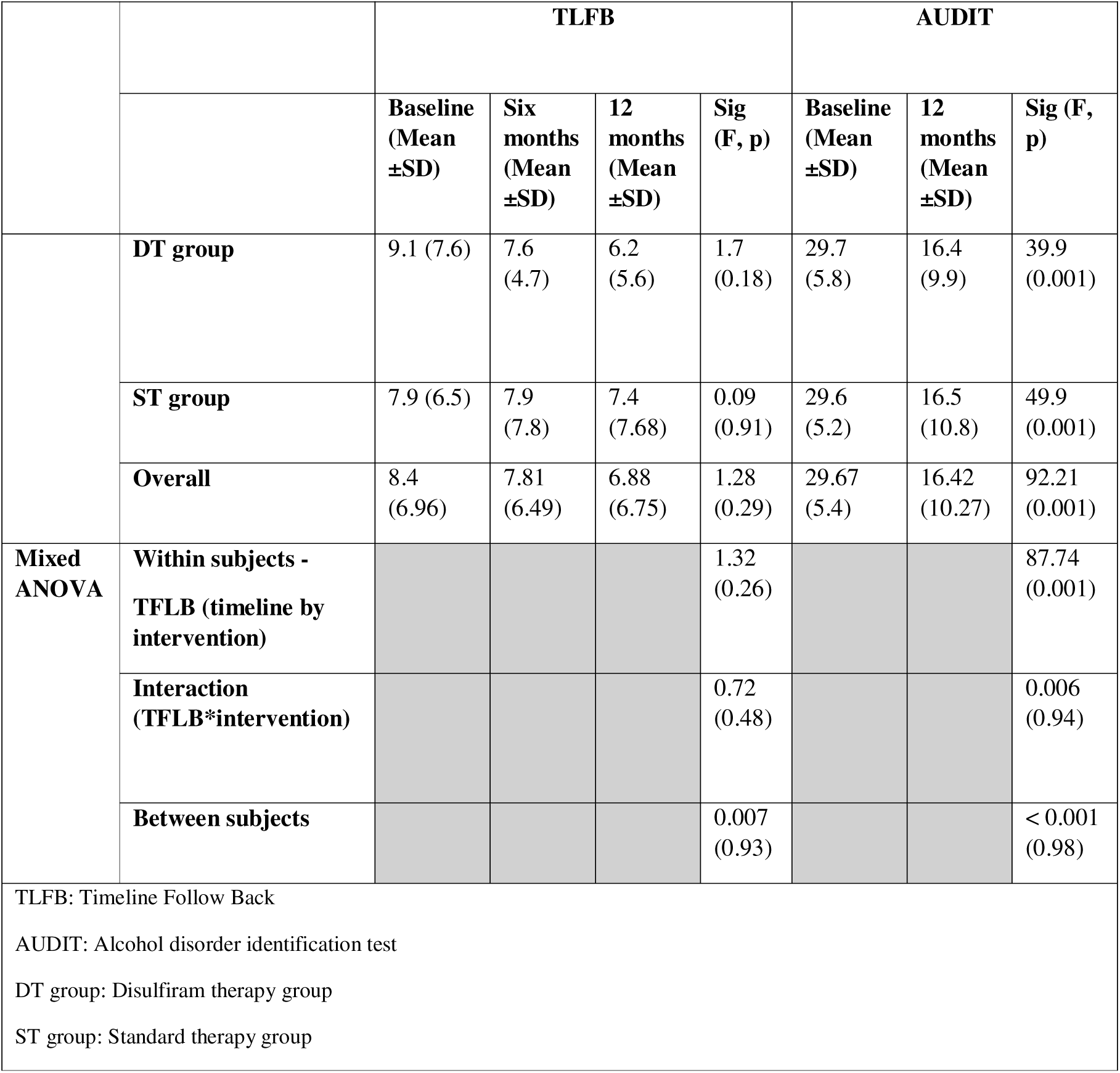
Analysis of TLFB and AUDIT scores over time.

|  |  | TLFB |  |  |  | AUDIT |  |  |
| --- | --- | --- | --- | --- | --- | --- | --- | --- |
|  |  | Baseline<br>(Mean<br>±SD) | Six<br>months<br>(Mean<br>±SD) | 12<br>months<br>(Mean<br>±SD) | Sig<br>(F, p) | Baseline<br>(Mean<br>±SD) | 12<br>months<br>(Mean<br>±SD) | Sig (F,<br>p) |
|  | **DT group** | 9.1 (7.6) | 7.6<br>(4.7) | 6.2<br>(5.6) | 1.7<br>(0.18) | 29.7<br>(5.8) | 16.4<br>(9.9) | 39.9<br>(0.001) |
|  | **ST group** | 7.9 (6.5) | 7.9<br>(7.8) | 7.4<br>(7.68) | 0.09<br>(0.91) | 29.6<br>(5.2) | 16.5<br>(10.8) | 49.9<br>(0.001) |
|  | **Overall** | 8.4<br>(6.96) | 7.81<br>(6.49) | 6.88<br>(6.75) | 1.28<br>(0.29) | 29.67<br>(5.4) | 16.42<br>(10.27) | 92.21<br>(0.001) |
| **Mixed ANOVA** | **Within subjects -**<br>**TFLB (timeline by**<br>**intervention)** |  |  |  | 1.32<br>(0.26) |  |  | 87.74<br>(0.001) |
|  | **Interaction**<br>**(TFLB\*intervention)** |  |  |  | 0.72<br>(0.48) |  |  | 0.006<br>(0.94) |
|  | **Between subjects** |  |  |  | 0.007<br>(0.93) |  |  | < 0.001<br>(0.98) |
| TLFB: Timeline Follow Back<br>AUDIT: Alcohol disorder identification test<br>DT group: Disulfiram therapy group<br>ST group: Standard therapy group |  |  |  |  |  |  |  |  |

Table 4 shows that all three biomarkers, ALT (F=4.70, p=0.012), γ-GT (F=38.6, p=0.001), and MCV (F=13.8, p=0.001), demonstrated statistically improvements from baseline to 12 months in DT group. While the improvement of ALT and MCV was modest, γ-GT showed a highly clinically significant improvement. The improvement seen in ALT (F=5.30, p=0.022), γ-GT (F=9.35, p=0.004), and MCV (F=21.90, p=0.001) was similarly significant in the ST group. Despite these robust within-group improvements, between-group comparisons revealed no significant difference in biomarker reduction (F=0.815, p=0.37), indicating equivalent efficacy of DT and ST on physiological measures of alcohol-related harm(Figure 3).

**Table 4:** Analysis of biomarkers over three time points.

| Table 4: Analysis of biomarkers over three time points |  |  |  |  |  |  |  |  |  |  |  |  |
| --- | --- | --- | --- | --- | --- | --- | --- | --- | --- | --- | --- | --- |
|  | ALT |  |  |  | γ-GT |  |  |  | MCV |  |  |  |
|  | Basel<br>ine<br>(Mea<br>n<br>±SD) | Six<br>mon<br>ths<br>(Me<br>an<br>±SD<br>) | 12<br>mon<br>ths<br>(Me<br>an<br>±SD<br>) | Sig<br>(F,<br>p) | Basel<br>ine<br>(Mea<br>n<br>±SD) | Six<br>mon<br>ths<br>(Me<br>an<br>±SD<br>) | 12<br>mon<br>ths<br>(Me<br>an<br>±SD<br>) | Sig<br>(F,<br>p) | Basel<br>ine<br>(Mea<br>n<br>±SD) | Six<br>mon<br>ths<br>(Me<br>an<br>±SD<br>) | 12<br>mon<br>ths<br>(Me<br>an<br>±SD<br>) | Sig<br>(F,<br>p) |
| DT group | 52.2<br>±<br>25.5 | 40.2<br>±<br>5.2 | 47.9<br>±<br>17.4 | 4.7<br>(0.0<br>12) | 174.1<br>±<br>77.7 | 93.3<br>±<br>40.9 | 53.8<br>±<br>12.9 | 38.6<br>(0.0<br>01) | 93.4<br>± 6.5 | 88.6<br>±<br>2.3 | 92.9<br>±<br>1.9 | 13.8<br>(0.0<br>01) |
| ST group | 60.4<br>±<br>47.1 | 40.6<br>±<br>6.1 | 44.7<br>±<br>12.8 | 5.3<br>(0.0<br>22) | 180.7<br>±<br>210.6 | 98.0<br>±<br>38.3 | 56.6<br>±<br>18.9 | 9.35<br>(0.0<br>04) | 94.7<br>± 4.5 | 88.9<br>±<br>4.6 | 92.9<br>±<br>3.1 | 21.9<br>(0.0<br>01) |
| Overall | 56.63<br>(38.4) | 40.5<br>(5.6) | 46.1<br>(14. | 8.85<br>(0.0 | 177.0<br>(161. | 96.6<br>2 | 55.5<br>8 | 27.1<br>3 | 94.18<br>(5.44 | 88.7<br>9 | 92.8<br>7 | 35.7<br>(0.0 |
|  | 2) | 9) | 9) | 02) | 69) | (39.57) | (16.4) | (0.001) | ) | (3.71) | (2.58) | 01) |
| **Within subjects -**<br>**(timeline by intervention)** |  |  |  | 8.25<br>(0.001) |  |  |  | 26.29<br>(0.001) |  |  |  | 34.56<br>(0.001) |
| **Interaction (category\*intervention)** |  |  |  | 1.1<br>(0.339) |  |  |  | 0.006<br>(0.945) |  |  |  | 0.469<br>(0.627) |
| **Between subjects** |  |  |  | 0.23<br>(0.62) |  |  |  | 0.14<br>(0.708) |  |  |  | 0.815<br>(0.37) |
| ALT: Alanine transaminase<br>γ-GT: γ-glutamyl transaminase<br>MCV: Mean corpuscular volume<br>DT group: Disulfiram therapy group<br>ST group: Standard therapy group |  |  |  |  |  |  |  |  |  |  |  |  |

Analysis of cognitive-affective-behavioural factors, as shown in Table 5, revealed distinct patterns of improvement across treatment groups. Both DT (F=53.12, p=0.001) and ST (F=40.2, p=0.001) groups demonstrated highly significant improvements in AARSU scores. However, direct comparison between groups showed no significant difference in efficacy (F=0.2, p=0.818). In contrast, BARSU, the behavioural component, showed differential outcomes between treatments. While the DT group’s improvement did not reach statistical significance (F=11.78, p=0.27), the ST group demonstrated a robust increase (F=28.70, p=0.001). Despite this apparent divergence, between-group analysis confirmed no statistically significant difference in treatment effects (F=0.37, p=0.69).

**Table 5:**
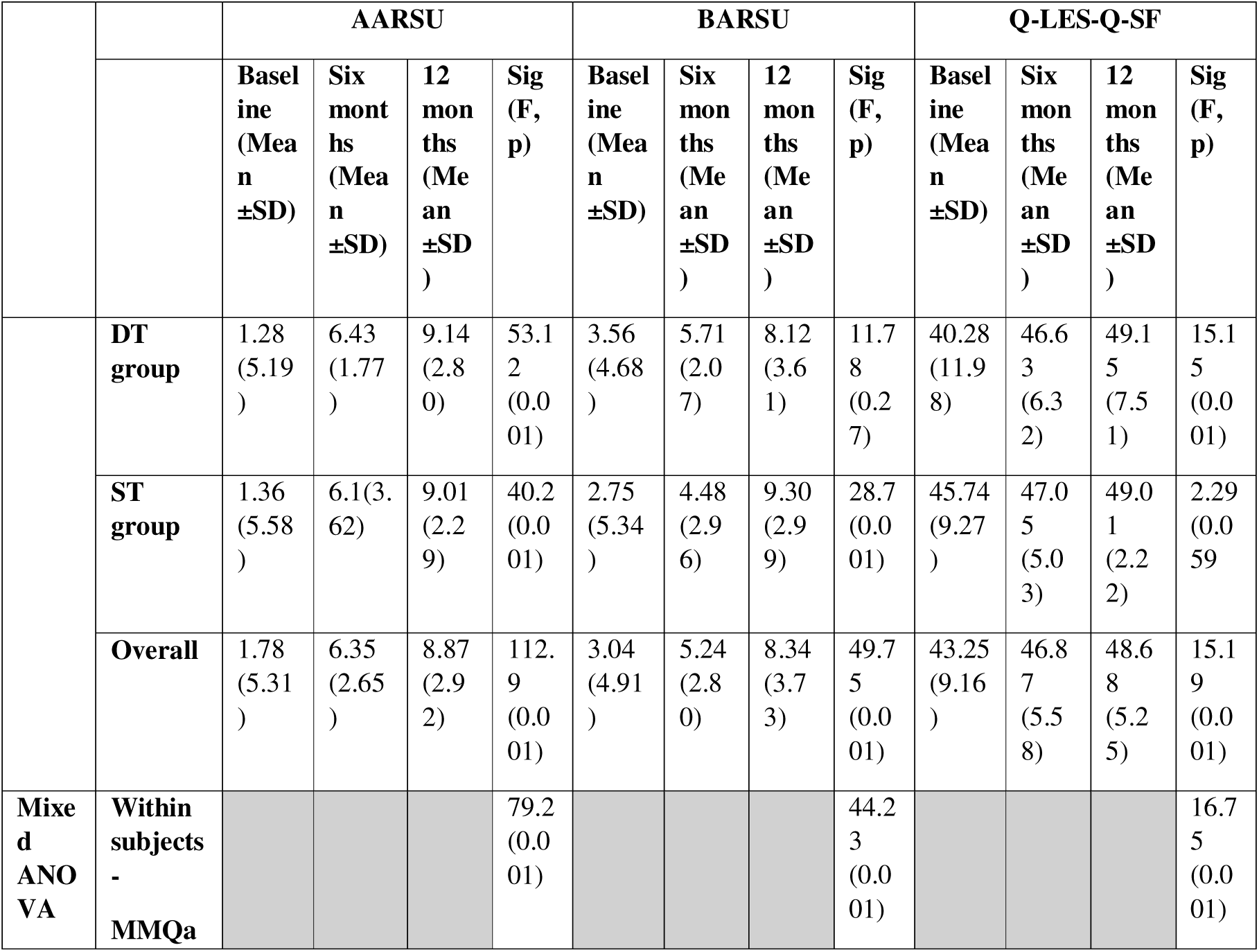
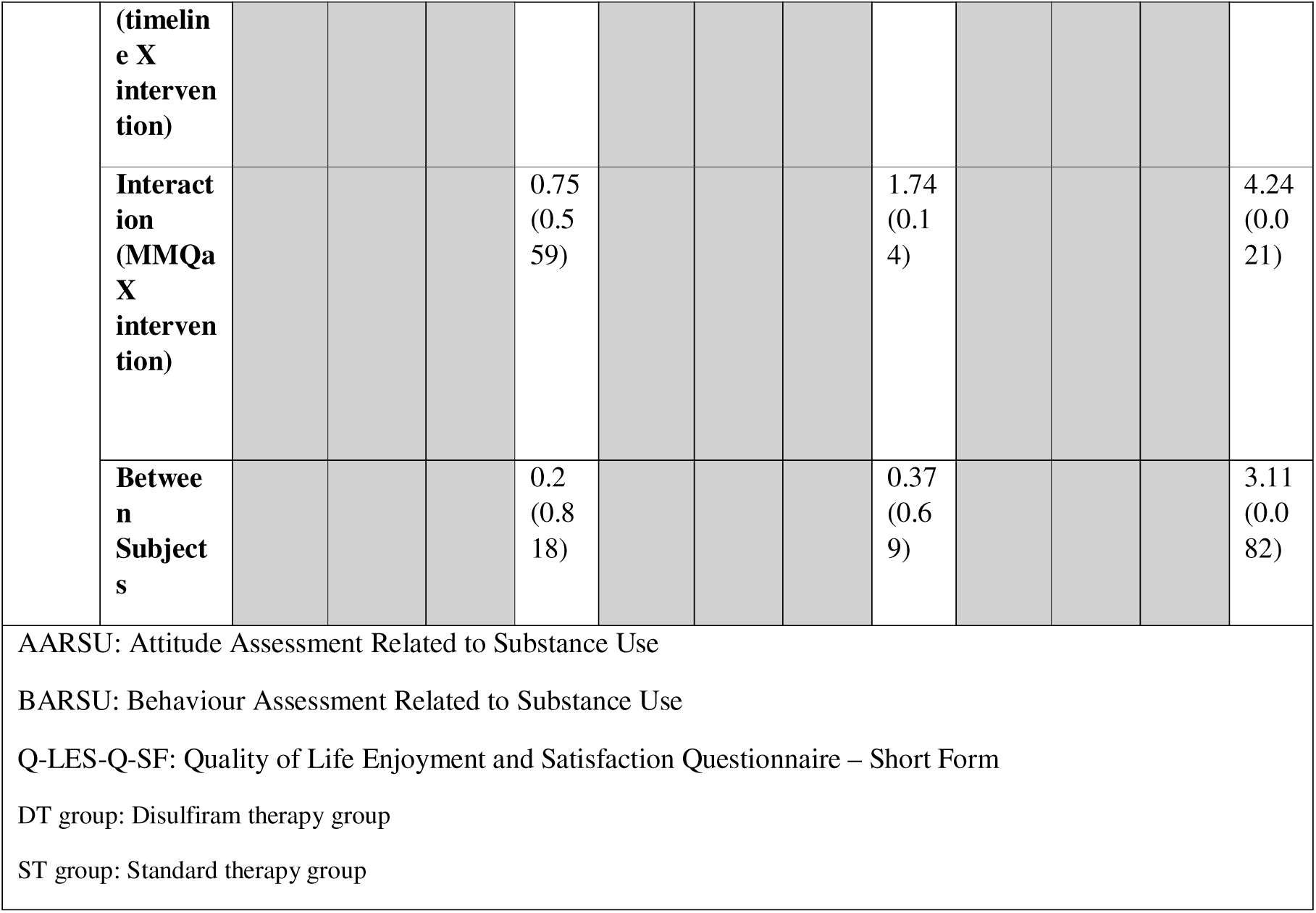
Analysis of cognitive-affective-behavioural factors and quality of life outcomes over time.

With respect to quality-of-life measures (Table 5), DT participants experienced a statistically significant enhancement in Q-LES-Q-SF scores (F=15.15, p=0.001), while ST participants showed no improvement (F=2.29, p=0.059). Importantly, direct comparison between treatments revealed no statistically significant advantage for either approach (F=3.11, p=0.082).

Treatment adherent patterns were varied in both groups. Missed clinic appointments were usually associated with discontinuation of disulfiram, sometimes with reinstatement of alcohol use, in the DT group. Resuming clinic follow up and disulfiram therapy was found to have been encouraged by the family.

### Impact of motivation and diagnosis at baseline

Analysis of baseline motivation, as shown in Table 6, revealed that treatment (either disulfiram or standard) reduced AUDIT scores in both high- and low-motivation groups (F = 82.3, p < 0.001).

**Table 6:**
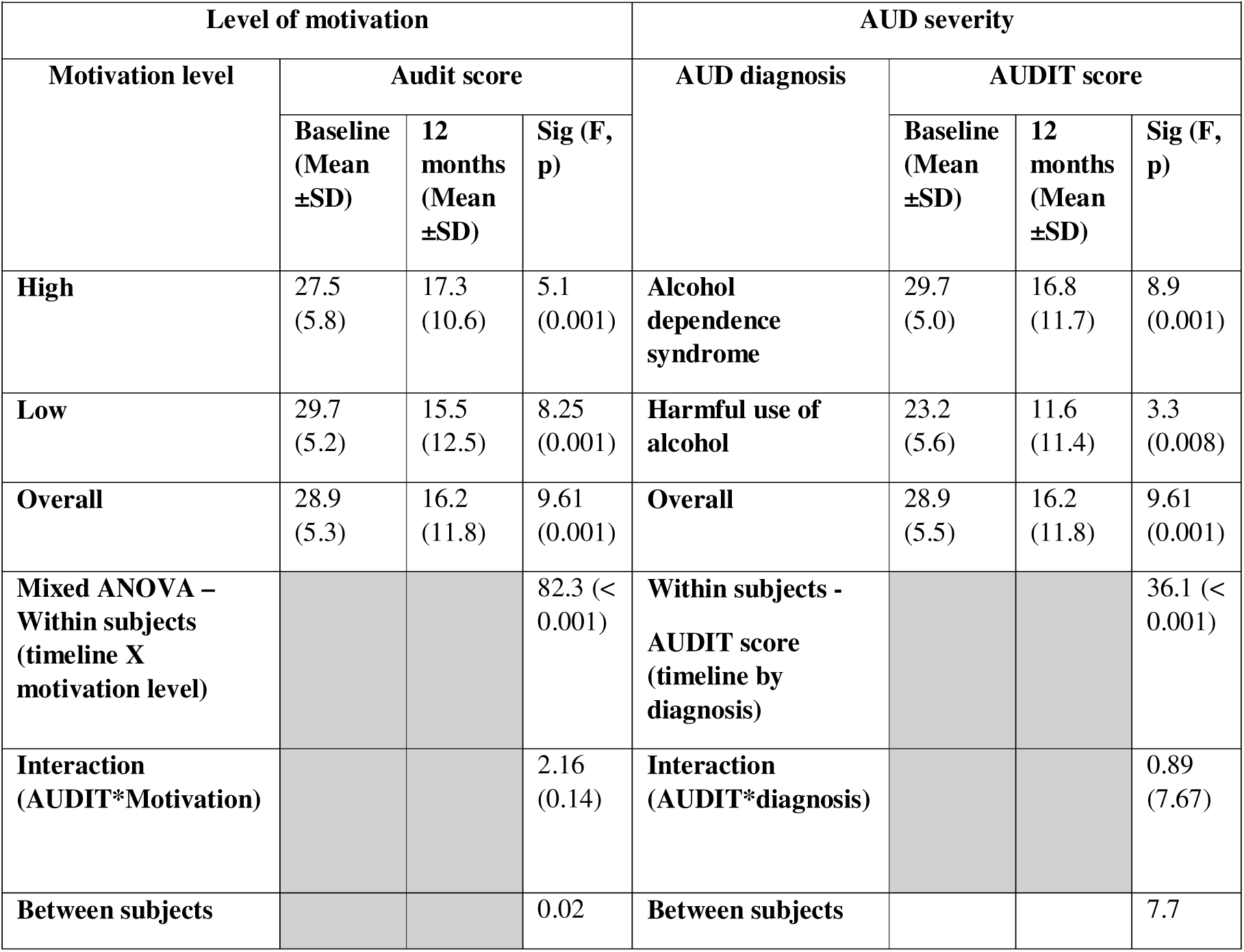
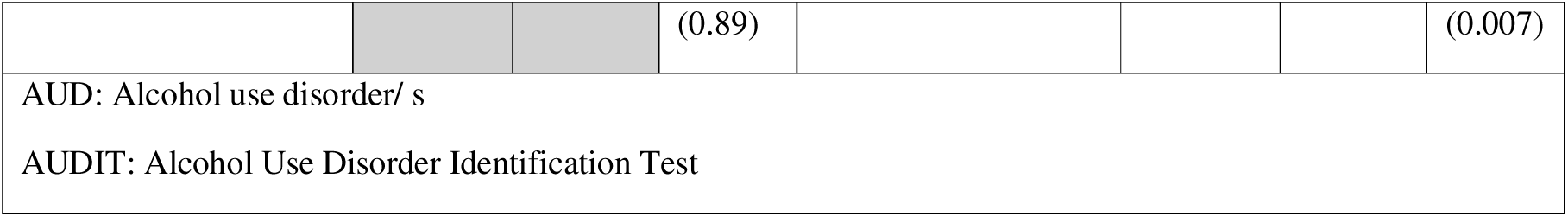
Analysis of baseline motivation level and AUD severity on subsequent treatment response.

However, the absence of any significant between-group difference (F = 0.20, p = 0.89) implies that level of motivation at baseline did not moderate treatment outcomes since both groups showed comparable gains. In contrast, baseline severity of diagnosis showed a direct moderating effect. While both alcohol dependence and harmful use participants significantly improved after treatment (F = 36.10, p < 0.001), difference in improvement between the two groups was significant (F = 7.70, p = 0.007). Those with a worse presentation at baseline, i.e., individuals with alcohol dependence versus those with harmful use of alcohol, demonstrated a greater improvement in AUDIT scores with either treatment(Figure 2).

**Figure 2.**
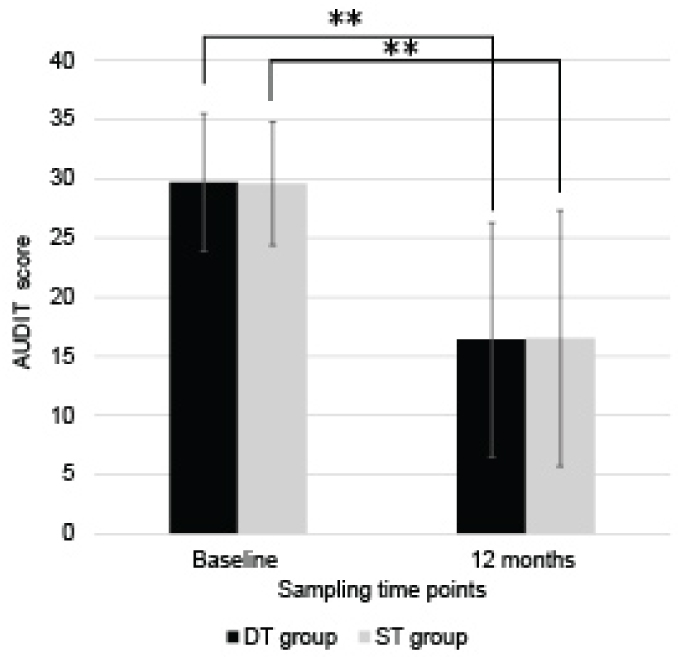
Mean AUDIT scores at baseline and 12-month follow-up in the disulfiram therapy (DT) and standard treatment (ST) groups. Significant reduction in AUDIT scores were observed within both groups over time (p < 0.01), with no significant group × time interaction. Error bars represent standard deviation. Ns P > 0.05, *P ≤ 0.05, ** P ≤ 0.01,

**Figure 3.**
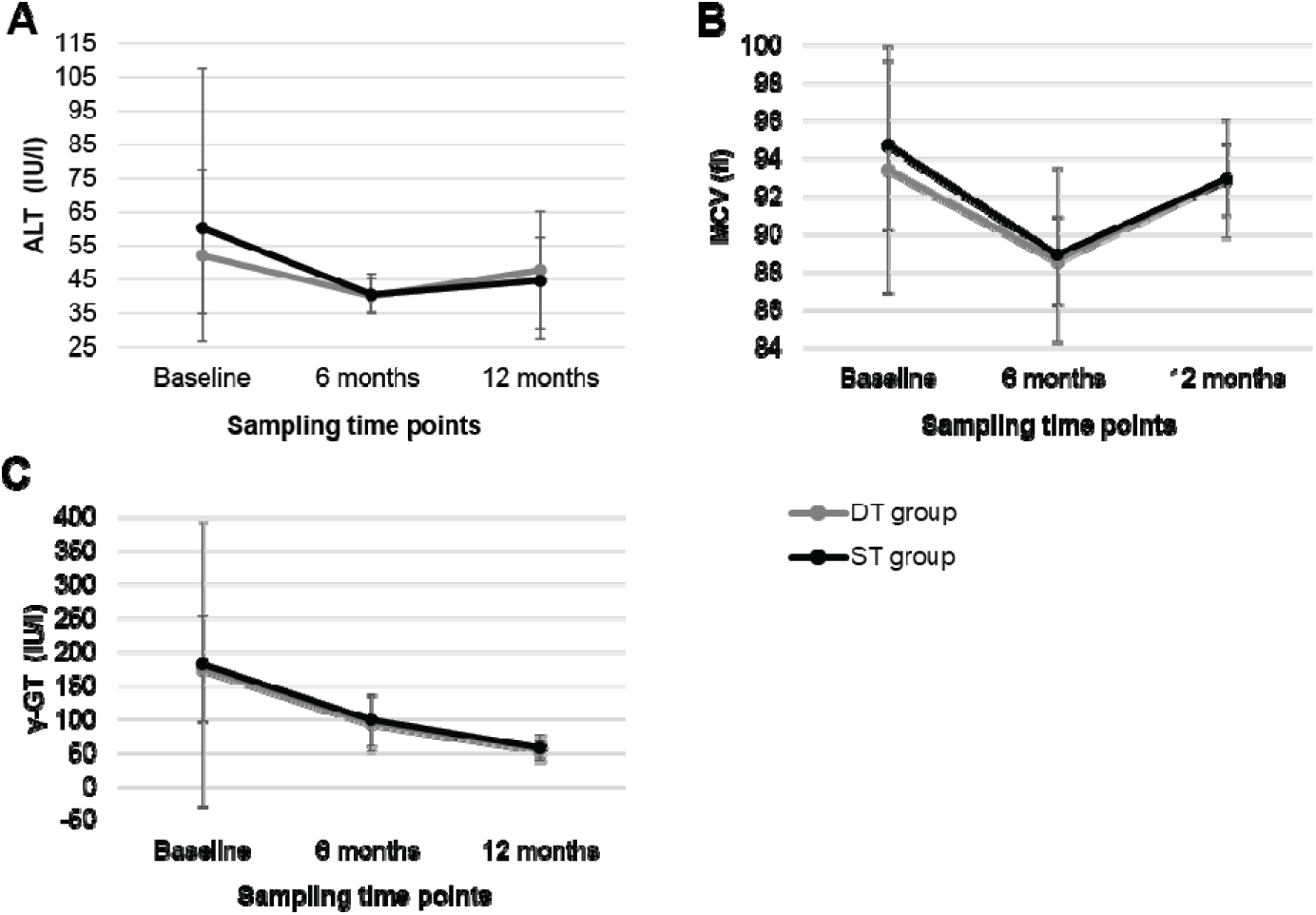
Mean levels of alcohol related biomarkers over time in the disulfiram therapy (DT) and standard treatment (ST) groups: (A) alanine aminotransferase (ALT), (B) mean corpuscular volume (MCV), and (C) gamma-glutamyl transferase (γ-GT). A significant within group improvement was observed along the study time period in both treatment groups while having no significant group × time interaction for any biomarker. Error bars represent standard deviation.

## Discussion

The main conclusion of this pragmatic RCT designed on a biopsychosocial foundation is that family-supervised disulfiram was as effective as the standard treatment in a real-world setting for patients with AUD. This is the first time that disulfiram supervised by family was shown to be as effective as the standard treatment for AUD in Sri Lanka. The participants of the DT group accepted involvement of family, which indicates that family-supervised disulfiram was culturally acceptable. However, they displayed variable and intermittent adherence to disulfiram therapy, and hence variable and intermittent abstinence from alcohol.

Intermittent use of disulfiram in patients having significant family support is a peculiar observation in this study, which highlights several important points: 1. All participants successfully engaged a family member or, sometimes, a friend as the supervisor, confirming the cultural acceptability of this approach in Sri Lanka. 2. This mirrors older studies of supervised disulfiram using informants, usually a family member from the developed world (8–11). 3. The pragmatic design of the trial allowed intermittent use of disulfiram reflecting its real-world implementation. 4. The family member was very likely to apply pressure to restart disulfiram once stopped. 5. Despite variable adherence without protocol-enforced reminders, supervised disulfiram still yielded significant improvements, although not prolonged total abstinence, which could have been the case if ETAT-RCT was a per-protocol trial.

Since there was no treatment matching in the trial, history of past treatment was not deemed relevant. All participants randomly allocated to the DT group were offered disulfiram irrespective of their treatment history. Hence, family-supervised disulfiram therapy in the ETAT-RCT, found to be as effective as the standard therapy, was essentially the first-line treatment option for the participants of the DT group within this clinical trial.

The null between-group differences may reflect our trial’s high pragmatism, which prioritised effectiveness (real-world performance) over efficacy (idealised conditions). A more controlled, per-protocol design might have amplified DT’s effects, particularly on alcohol consumption, as shown before (4,7,17). The ETAT-RCT was expressly designed as a pragmatic trial, fully meeting seven of Ford and Norie’s nine criteria for pragmatic clinical research (22). This design prioritised real-world applicability through recruitment of a representative clinical population, flexible adherence protocols mirroring routine practice, and outcome measures tied to clinical decision-making.

The high degree of pragmatism in our methods suggests our findings largely reflect not just *efficacy* (performance under ideal conditions), but *effectiveness* (performance in typical clinical settings). Hence, our findings underscore that in routine practice, supervised disulfiram, even with fluctuating adherence, may be as effective as standard care. This has critical implications for resource allocation in low- and middle-income countries like Sri Lanka, where rolling out supervised disulfiram as a first-line treatment in AUD is likely to be more appropriate and feasible than other psychological therapies or costlier pharmacotherapy. It is acknowledged that disulfiram does carry certain risks over other treatments, but a full risk-benefit evaluation is outside the scope of this paper.

Treatment guidelines emphasise that disulfiram to be reserved for highly motivated patients with more severe forms of AUD who are aiming for abstinence, effectively removing it from the first-line treatment options (16)(18,19). The American Psychiatric Association Practice Guideline includes another prerequisite – having experienced intolerance or lack of response to naltrexone and acamprosate (19). However, our findings suggest otherwise making a clear case for its use as a first-line treatment in AUD. Furthermore, motivation level (high vs. low) did not moderate outcomes in our study. AUD severity (dependence vs. harmful use) did have an effect, where patients with more severe AUD at the baseline benefiting more than the others irrespective of the type of treatment.

Our findings indicate that benefits of supervised disulfiram as a first line treatment in AUD in Sri Lanka may extend beyond self-selected, highly motivated abstinence-focused subgroups. Our sample was predominantly Sinhalese (74.6%, n=53) and Buddhist (70.4%, n=50), mirroring the demographic profile of the local patient population.

The instruments used in the ETAT-RCT to measure progress and treatment effectiveness are multifaceted. In both disulfiram and standard treatment groups, the instruments were able to demonstrate significant pre- to post-treatment improvements in pathological alcohol use (AUDIT scores, biomarkers), positive shifts in cognitive-affective-behavioural factors (AARSU/BARSU), and enhanced quality of life (Q-LES-Q-SF). However, the TLFB score showed only modest, non-significant improvement—a finding that warrants careful consideration given the tool’s unvalidated translation in this population (see *Limitations*).

### Generalisability and applicability

The findings of the ETAT-RCT are most applicable in low- and middle-income countries where public-sector addiction services operate with limited resources. The trial reflects outcomes expected in real world delivery of health service rather than idealised research settings, by conducting the study in an unmodified public hospital setting without additional staffing, structured adherence monitoring, or enhanced follow-up. It is interesting to note that 88.9% of those screened were found to be eligible and only less than 0.02% declined to participate, which indicate wide objective and subjective applicability of the treatment interventions offered in ETAT-RCT. All participants allocated to DT group were able to nominate a family member or trusted individual to supervise treatment, suggesting transferability to settings with comparable family and community cohesion similar to this Sri Lankan study population. Contrastingly, applicability may be limited in contexts where family and community involvement are culturally constrained.

The pragmatic design further enhances relevance to health systems where resources are constrained. Disulfiram was delivered at minimal cost, and no strategies were introduced to enhance the adherence, allowing natural variability in clinic attendance and adherence to treatments. This resonates with routine public sector practice and indicates that effectiveness does not depend on intensive monitoring or specialised infrastructure. Generalisability is shaped by the male-only sample, reflecting local epidemiology and help seeking patterns.

Overall, the ETAT-RCT supports family supervised disulfiram as a feasible first line option within comparable public sector health systems in populations with high family and community cohesion.

## Limitations

Due to the nature of the natural presentation to treatment services at public hospitals, but not due to a flaw of the design of the study, the participants were exclusively male limiting the generalisability of its findings. Even though this limits extrapolation to women, it strengthens applicability to similar clinical settings where treatment populations are predominantly male. The diverse nature of the delivery of the treatments prevented ETAT-RCT being designed as a double-blind RCT, which may have introduced bias into self-reported outcomes. The unvalidated Sinhala TLFB translation limits interpretation of alcohol-consumption outcomes. While its interviewer-administered format mitigated language barriers, the tool’s reliance on standard-drink recall in a culturally novel context may have introduced measurement error. The treatment heterogeneity in the ST group (optional naltrexone, variable psychotherapeutic input), despite reflecting real-world practice, may have obscured between-group differences. The 12-month assessment captures medium-term outcomes but cannot speak to long-term relapse rates or sustainability of observed improvements. ETAT-RCT lacked the capacity to measure the extent of periods of abstinence rendering deeper evaluation of intermittent use of disulfiram with sporadic periods of total abstinence in the DT group constrained.

## Conclusions

Disulfiram therapy supervised by a nominated family member constitute an acceptable and a practical first line treatment intervention in AUD. The effectiveness of supervised disulfiram therapy is unaffected by the severity of the AUD or the level of motivation. These findings advocate for patient-centred AUD treatment selection, where factors like family support and quality-of-life goals, not just sustained abstinence but guide clinical decision-making.

## Trial registration

This RCT was registered in the Sri Lanka Clinical Trials Registry (https://www.slctr.lk/) under the name “Efficacy of two treatment strategies for chronic alcohol use disorders in comparison to that currently practiced at the Psychiatric Unit of the University of Colombo - a RCT” on 19 September 2014 (SLCTR/2014/021).

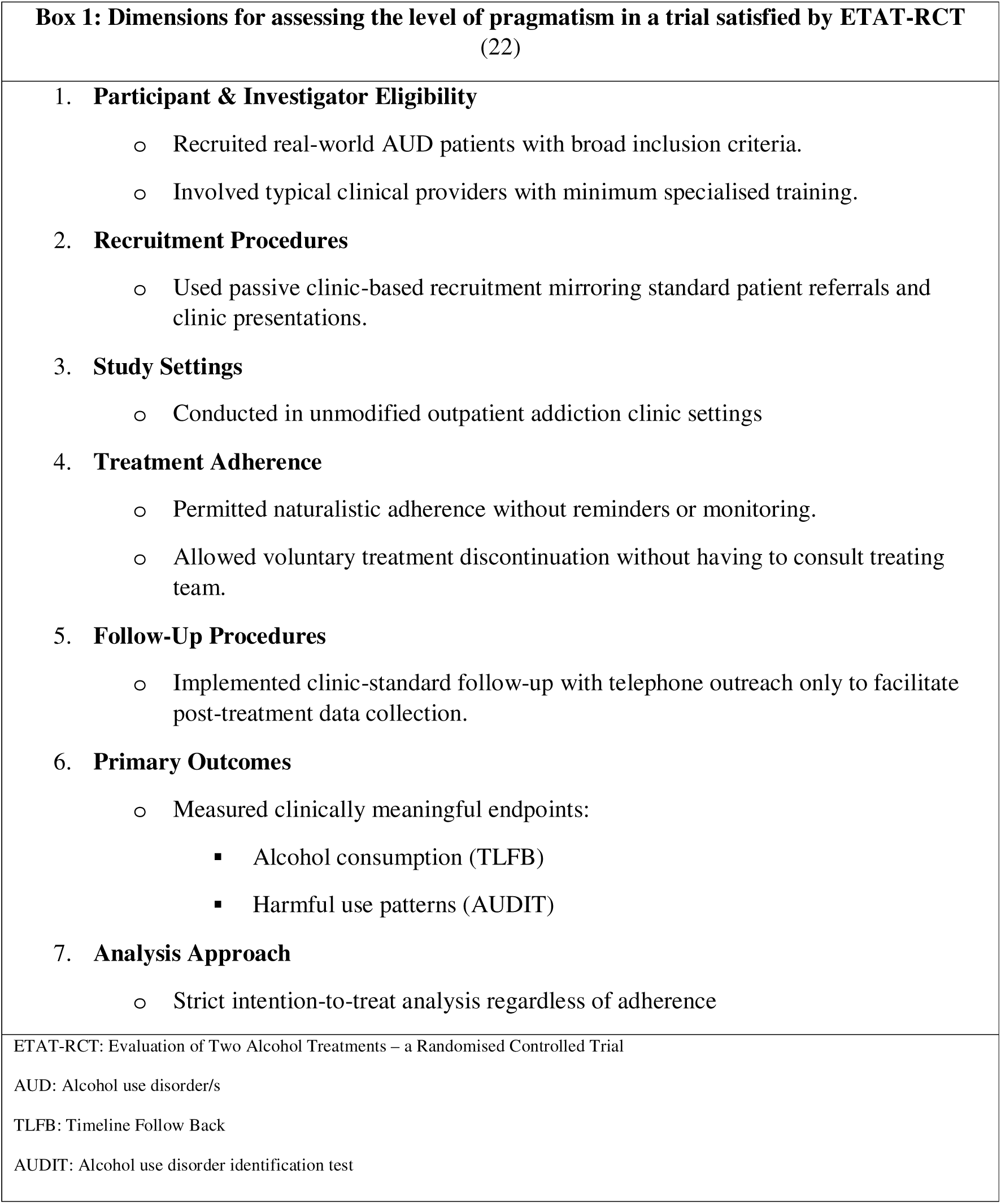

| **Box 1: Dimensions for assessing the level of pragmatism in a trial satisfied by ETAT-RCT**<br>(22) |
| --- |
| 1. **Participant & Investigator Eligibility**<br>○ Recruited real-world AUD patients with broad inclusion criteria.<br>○ Involved typical clinical providers with minimum specialised training.<br>2. **Recruitment Procedures**<br>○ Used passive clinic-based recruitment mirroring standard patient referrals and clinic presentations.<br>3. **Study Settings**<br>○ Conducted in unmodified outpatient addiction clinic settings<br>4. **Treatment Adherence**<br>○ Permitted naturalistic adherence without reminders or monitoring.<br>○ Allowed voluntary treatment discontinuation without having to consult treating team.<br>5. **Follow-Up Procedures**<br>○ Implemented clinic-standard follow-up with telephone outreach only to facilitate post-treatment data collection.<br>6. **Primary Outcomes**<br>○ Measured clinically meaningful endpoints:<br>▪ Alcohol consumption (TLFB)<br>▪ Harmful use patterns (AUDIT)<br>7. **Analysis Approach**<br>○ Strict intention-to-treat analysis regardless of adherence |
| ETAT-RCT: Evaluation of Two Alcohol Treatments – a Randomised Controlled Trial<br>AUD: Alcohol use disorder/s<br>TLFB: Timeline Follow Back<br>AUDIT: Alcohol use disorder identification test |

## Data Availability

All data produced in the present study are available upon reasonable request to the authors

## Acknowledgements

The authors are grateful to the supervisors of the ETAT-RCT Raveen Hanwella and The Late Robert Mann for their guidance and support, and to Nalika Gunwardena for her input in upgrading the methodology. The contribution from the IPAT team in delivery of therapy for the PT group is deeply appreciated. Staff of UPU, especially Kasturi Sayakkara, are fondly remembered for their unwavering support.

## Author contributions

MR conceived, designed, and carried out the trial as the principal investigator and compiled the initial draft manuscript. PC carried out the statistical analysis. DP and PR contributed to upgrade the quality of the manuscript. All authors had a key role in revising the manuscript and approved the final version.

## Reporting statement

The trial followed CONSORT 2010 guidelines

## Patient and public involvement

Patients and or public were not involved in the design, conduct, reporting, or dissemination plans of this research. Refer to the Methods section for further details.

## Data availability statement

Data available for access upon request from the corresponding author, subject to ERC recommendations and approval.

## Provenance and peer review

Externally peer reviewed.

---

**Figure 1.**
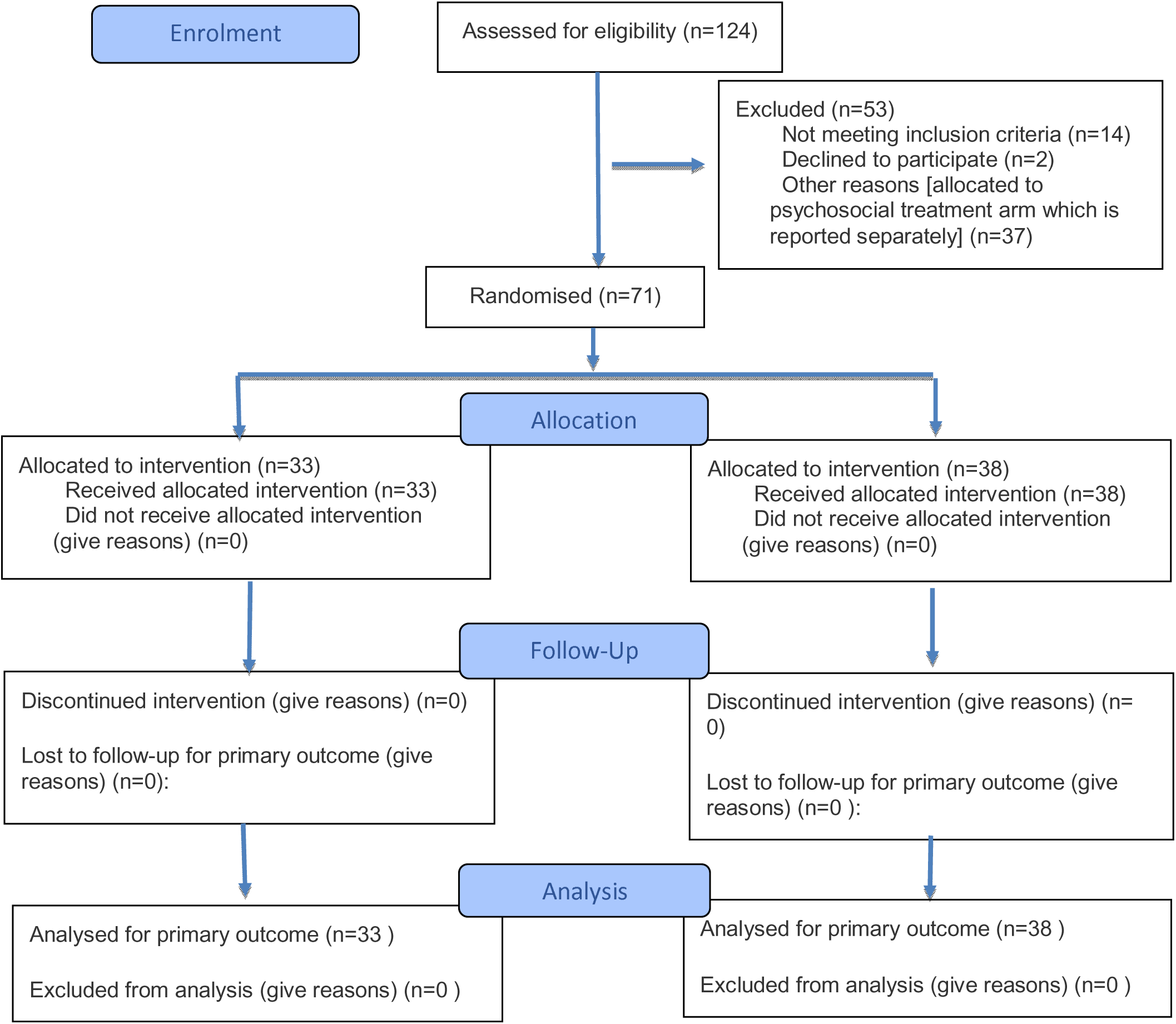
CONSORT 2025 Flow Diagram. Flow diagram of the progress through the phases of a randomised trial of two groups (that is, enrolment, intervention allocation, follow-up, and data analysis)

